# Early Treatment, Inflammation and Post-COVID Conditions

**DOI:** 10.1101/2023.02.13.23285855

**Authors:** Kelly A. Gebo, Sonya L. Heath, Yuriko Fukuta, Xianming Zhu, Sheriza Baksh, Alison G. Abraham, Feben Habtehyimer, David Shade, Jessica Ruff, Malathi Ram, Oliver Laeyendecker, Reinaldo E. Fernandez, Eshan U. Patel, Owen R. Baker, Shmuel Shoham, Edward R. Cachay, Judith S. Currier, Jonathan M. Gerber, Barry Meisenberg, Donald N. Forthal, Laura L. Hammitt, Moises A. Huaman, Adam Levine, Giselle S. Mosnaim, Bela Patel, James H. Paxton, Jay S. Raval, Catherine G. Sutcliffe, Shweta Anjan, Thomas Gniadek, Seble Kassaye, Janis E. Blair, Karen Lane, Nichol A. McBee, Amy L. Gawad, Piyali Das, Sabra L. Klein, Andrew Pekosz, Arturo Casadevall, Evan M. Bloch, Daniel Hanley, Aaron A.R. Tobian, David J. Sullivan, the CSSC-004 Consortium

## Abstract

**Background:** Post-COVID conditions (PCC) are common and have significant morbidity. Risk factors for PCC include advancing age, female sex, obesity, and diabetes mellitus. Little is known about early treatment, inflammation, and PCC.

**Methods:** Among 883 individuals with confirmed SARS-CoV-2 infection participating in a randomized trial of CCP vs. control plasma with available biospecimens and symptom data, the association between early COVID treatment, cytokine levels and PCC was evaluated. Cytokine and chemokine levels were assessed at baseline, day 14 and day 90 using a multiplexed sandwich immuosassay (Mesoscale Discovery). Presence of any self-reported PCC symptoms was assessed at day 90. Associations between COVID treatment, cytokine levels and PCC were examined using multivariate logistic regression models.

**Results:** One-third of the 882 participants had day 90 PCC symptoms, with fatigue (14.5%) and loss of smell (14.5%) being most common. Cytokine levels decreased from baseline to day 90. In a multivariable analysis including diabetes, body mass index, race, and vaccine status, female sex (adjusted odds ratio[AOR]=2.70[1.93-3.81]), older age (AOR=1.32[1.17-1.50]), and elevated baseline levels of IL-6 (AOR=1.59[1.02-2.47]) were associated with development of PCC. There was a trend for decreased PCC in those with early CCP treatment (≤5 days after symptom onset) compared to late CCP treatment.

**Conclusion:** Increased IL-6 levels were associated with the development of PCC and there was a trend for decreased PCC with early CCP treatment in this predominately unvaccinated population. Future treatment studies should evaluate the effect of early treatment and anti-IL-6 therapies on PCC development.

**Summary:** Increased IL-6 levels were associated with the development of Post-COVID Conditions (PCC) and there was a trend for decreased PCC with early COVID convalescent plasma treatment in this predominately unvaccinated population.

## INTRODUCTION

Severe Acute Respiratory Syndrome Coronavirus 2 (SARS-CoV-2) has infected more than 600 million people worldwide and more than 97 million people in the United States [1]. Many survivors of coronavirus disease 2019 (COVID-19) report long-term health effects including persistent symptoms, the development of new comorbidities and exacerbation of underlying pre-existing conditions. The medical community has formally recognized the long-term health effects of COVID-19 by establishing a diagnosis of post-COVID conditions (PCC). The World Health Organization (WHO) [2] and US Centers for Disease Control and Prevention (CDC) [3] differ in the definition of the timeline of this diagnosis, but both require that patients have sustained health conditions after resolution of acute COVID-19. According to the WHO, the diagnosis is a history of COVID-19 at least three months prior who have had symptoms for at least two months that an alternative diagnosis cannot explain [2]. The CDC defines PCC as symptoms persisting beyond four weeks after acute infection [4].

Although the true incidence of PCC is unknown due to differences in study populations and methodologies, its clinical impact is substantial. Recent data suggest that the prevalence is approximately 20%, with the most commonly reported symptoms being fatigue, shortness of breath, and cognitive dysfunction [5] with higher rates among those >65 years of age [6]. Previous studies have demonstrated several host factors that are associated with the development of PCC, including the severity of acute COVID-19, female sex [7-9], advanced age [6, 10, 11], being unvaccinated [11, 12], and presence of pre-COVID comorbidities.

Early treatment for COVID-19 is recommended to prevent hospitalization and death. While a recent observational study has suggested decreased PCC with early treatment with nirmatrelvir [13], the relationship between early effective COVID therapy and the development of PCC has been unclear. In addition to demographic factors and co-morbidities, inflammation appears to play a critical role in COVID-19 prognosis. Individuals with elevated levels of IL-6 and C-reactive protein are more likely to have more severe disease and an increased risk of hospitalization during the acute phase [14-18] particularly among males [19]. Few studies have evaluated whether cytokines are associated with PCC [20-26] and the limited data demonstrate inconsistent results. Some studies have shown an association between PCC and IL-6 [20, 22], while others have not [24, 25]. This is likely due to study limitations, including small sample sizes, lack of prospective monitoring of symptoms and sample collection, and inconsistent timing in the disease course in relation to cytokine evaluation.

To investigate these relationships, we analyzed prospectively collected data from participants enrolled in a randomized outpatient treatment trial of COVID-19 convalescent plasma (CCP) to identify factors associated with the development of PCC, including early treatment and biomarkers measured early in the course of disease.

## METHODS

### PARTICIPANTS

The Convalescent Plasma to Limit SARS-CoV-2 Associated Complications (CSSC-004) trial was a double-blind, multi-center, randomized, controlled trial investigating the use of CCP for the prevention of hospitalization among outpatients when compared to control plasma, as previously described [27]. The trial recruited 1225 symptomatic, adult outpatients with acute SARS-CoV-2 infection from June 3, 2020 to October 1, 2021, at 23 sites. Of these, 1181 participants received either CCP or control plasma transfusion. Trial participants were transfused within 9 days of symptom onset. Follow-up clinic visits were conducted at days 14, 28, and 90 post-transfusion. For this analysis, we restricted the study sample to those with blood drawn at screening, day 14, and day 90, and complete symptom data on day 90. Subsequently we excluded 21 screening samples, 23 day 14 samples, and 26 day 90 samples due to inadequate sample volume, laboratory error in cytokine measurement or missing results leading to exclusion of the sample. These lost samples were from different individuals at each time point; so these individuals were included in the analyses at other time points, when data was available.

#### Ethical Approvals

All study activities were approved by the Johns Hopkins University single Institutional Review Board, the Navajo Nation Human Research Review Board, the Indian Health Service National Institutional Review Board, and the Human Research Protection Office of the United States Department of Defense. All study activities followed the Declaration of Helsinki, the Good Clinical Practice guidelines of the International Conference on Harmonization, and all applicable regulatory requirements. Written informed consent was obtained from all study participants.

### OUTCOMES

The primary outcome was PCC, which was defined as the presence of any self-reported symptom (cough, fatigue, shortness of breath, headache, neurologic changes, loss of taste, loss of smell, nausea, vomiting, diarrhea, runny/stuffy nose, myalgias, sore throat, chills, fever, or skin manifestations) at the 90-day visit. All CSSC-004 trial participants were asked about the presence and severity of symptoms at days 0, 1, 3, 5, 7, 10, 14, 28 and 90 through a structured, self-report form administered via phone (days 1, 3, 5, 7, and 10) and at in-person visits (days 0, 14, 28 and 90).

#### Variable Definition

Age was modeled as a continuous variable in regressions. Sex was defined as biologic sex assigned at birth (male/female). Race (Black, American Indian/Alaskan Native [AI/AN], white, multiple, or other race) and ethnicity (Hispanic, non-Hispanic) were self-reported at enrollment. Body mass index (BMI) was calculated using height and weight at the screening visit. Calendar time was used as a surrogate for SARS-CoV-2 variant (transfusions prior to June 15, 2021 were classified as occurring during the pre-Alpha/Alpha wave, and those from June 15, 2021 to October 1, 2021 were classified as occurring during the Delta wave). Baseline comorbidities were self-reported at enrollment. Treatment was categorized into receipt of control plasma, early receipt <u>(<</u>5 days post symptom onset), and late receipt of CCP (> 5 days post symptom onset) – a timing threshold consistent with the original trial analysis [27]. Vaccination status was defined as fully vaccinated (i.e., more than 2 weeks from the second mRNA vaccine or two weeks from the first vector vaccine), partially vaccinated (i.e., receipt of one mRNA vaccine or less than two weeks from a vector vaccine) or unvaccinated.

#### Cytokine Measurement

Blood was collected in ethylenediaminetetraacetic acid (EDTA) tubes, and plasma was separated and isolated within 2-8 hours after collection by centrifugation and stored at –80°C until cytokines were measured.

Custom multiplexed sandwich immunoassays using MULTI-ARRAY electrochemiluminescence detection technology (MesoScale Discovery, Gaithersburg, MD) were used for the quantitative evaluation of 21 different human cytokine and chemokine analytes in plasma samples following the manufacturer’s instructions. The cytokines and chemokines were selected based on previous SARS-CoV-2 studies [28-30]. Analytes were considered “detectable” if both runs of each sample had a signal greater than the analyte- and plate-specific lower limit of detection (i.e., 2.5 standard deviations of the plate-specific blank). Analyte concentrations (pg/mL) from both runs of each analyte were averaged for analysis. Cytokine values below the lower limit of detection or above the upper limit of detection were estimated by the multiplex assay using extrapolation from the standard curve. Values above or below the fit curve range were reported missing by the assay.

### STATISTICAL ANALYSIS

Values that were not reported because they were above or below the fit curve were imputed using a single stochastic draw from the tail of a truncated log-normal distribution fit to the detectable values of each cytokine. For those below fit curve range, values were randomly drawn from the extrapolated lower tails (from 0 to the minimum available value) of the distribution for each analyte. For those above fit curve range, values were randomly drawn from the extrapolated upper tails (from the maximum available value to 10 times the available maximum) of the distribution for each analyte. Cytokine values were log_10_ transformed, and the outliers (*Q*3 + 1.5 × *interquartile range or Q*1 - 1.5 × *interquartile range*) from each visit were excluded to avoid potential lab error.

Two-tailed χ^2^ tests were performed to compare the baseline characteristics between treatment arms and PCC groups. Fischer’s exact test was used to compare across race categories due to a small number of participants in some racial groups. Spaghetti plots were produced to evaluate the trajectory of each analyte from screening to day 14 to day 90. Wilcoxon rank sum tests were used to compare the cytokine levels at screening between non-PCC and PCC groups. Similar to our previous treatment analyses and consistent with previously established risk factors for PCC, logistic regressions adjusted for the baseline covariates including age (continuous), sex (male vs. female), race (white vs. non-white), BMI (<30 vs. ≥30), vaccine status (partially or fully vs. none), diabetes (no vs. yes) were used to evaluate the association between each cytokine (log_10_ transformed values) and PCC. Benjamini-Hochberg correction with a false discovery rate of 0.05 was conducted to control for multiple comparisons across all the cytokines. Univariate and multivariate logistic regressions were modeled to estimate the potential association of baseline characteristics, IL-6, and CCP on the impact of PCC. Missing data on covariates were handled using available case method to include all cases where the variable(s) of interested were observed for each model. All analysis were performed in R 4.2.1 (R Core Team, Vienna, Austria).

## RESULTS

### STUDY POPULATION

Of 1181 transfused trial participants, 589 were randomized to receive control plasma and 592 were randomized to receive CCP, with 257 (43.4%) receiving CCP within 5 days of symptom onset (**Figure 1**). A total of 1061 participants returned for day 90 visits, of whom 882 had biorepository plasma samples and symptom data at baseline, day 14 and day 90. There were no significant differences in baseline characteristics, comorbidities, or COVID-19 vaccine status among those included in this study sampling compared to the entire clinical trial population (**Supplemental Table 1**).

**Figure 1.**
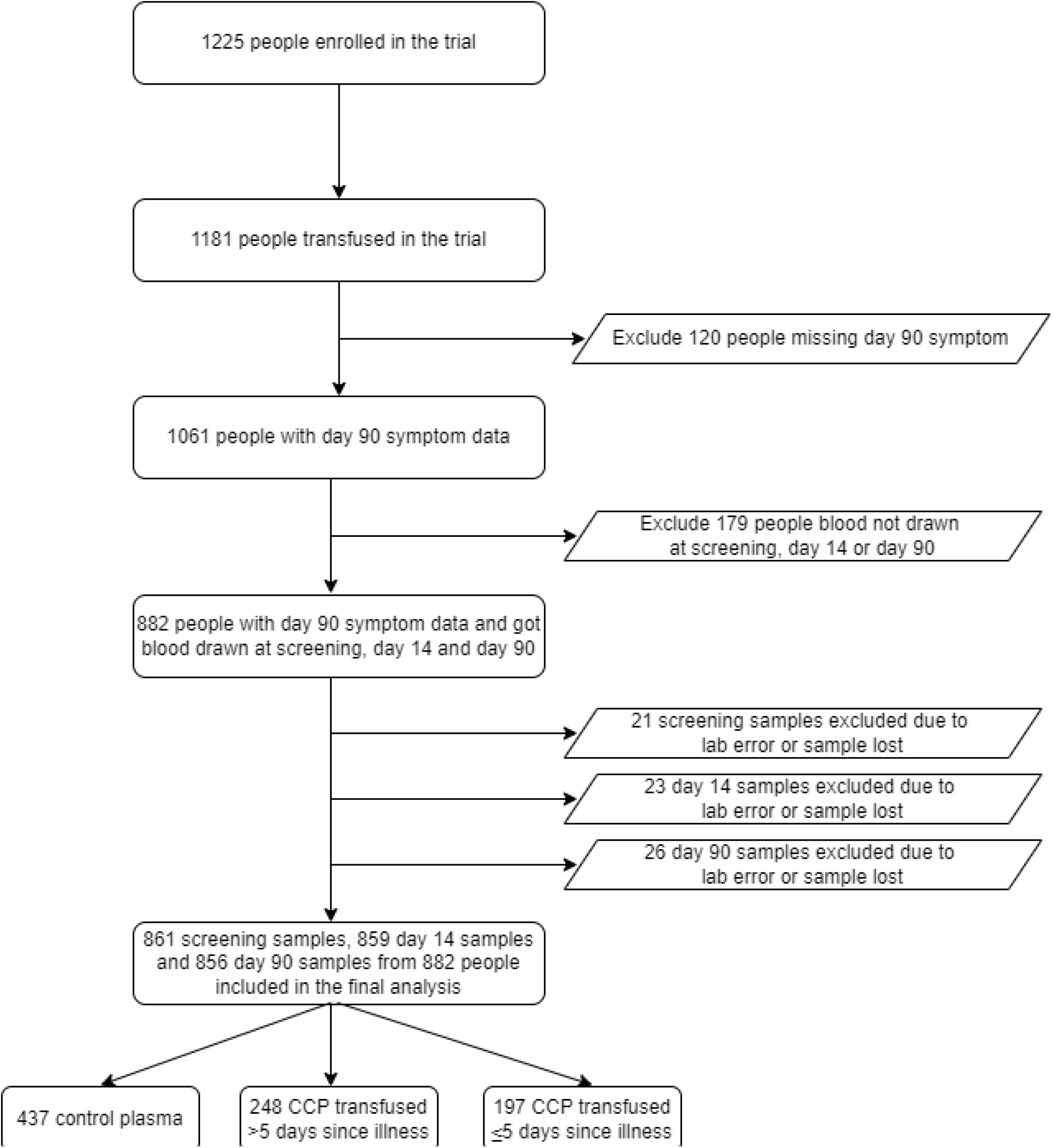
Study population.

The median age of the 882 subjects was 43 years, with 299 (33.9%) ≥ 50 years. Five hundred six (57.4%) female and 116 Black (13.2%) participants were included. There were no differences by trial treatment group (**Table 1**) or between those with and without PCC symptom data (**Supplemental Tables 2 & 3**). Demographic factors were also similar among those included in this analysis compared to the entire trial population (**Supplemental Table 1**). There were 590 (66.9%) individuals without PCC and 292 (33.1%) individuals with PCC at day 90 (**Table 2**). The most common reported symptoms were fatigue (14.5%), anosmia (14.5%), and ageusia (10.0%) (**Supplemental Table 4**).

**Table 1.**
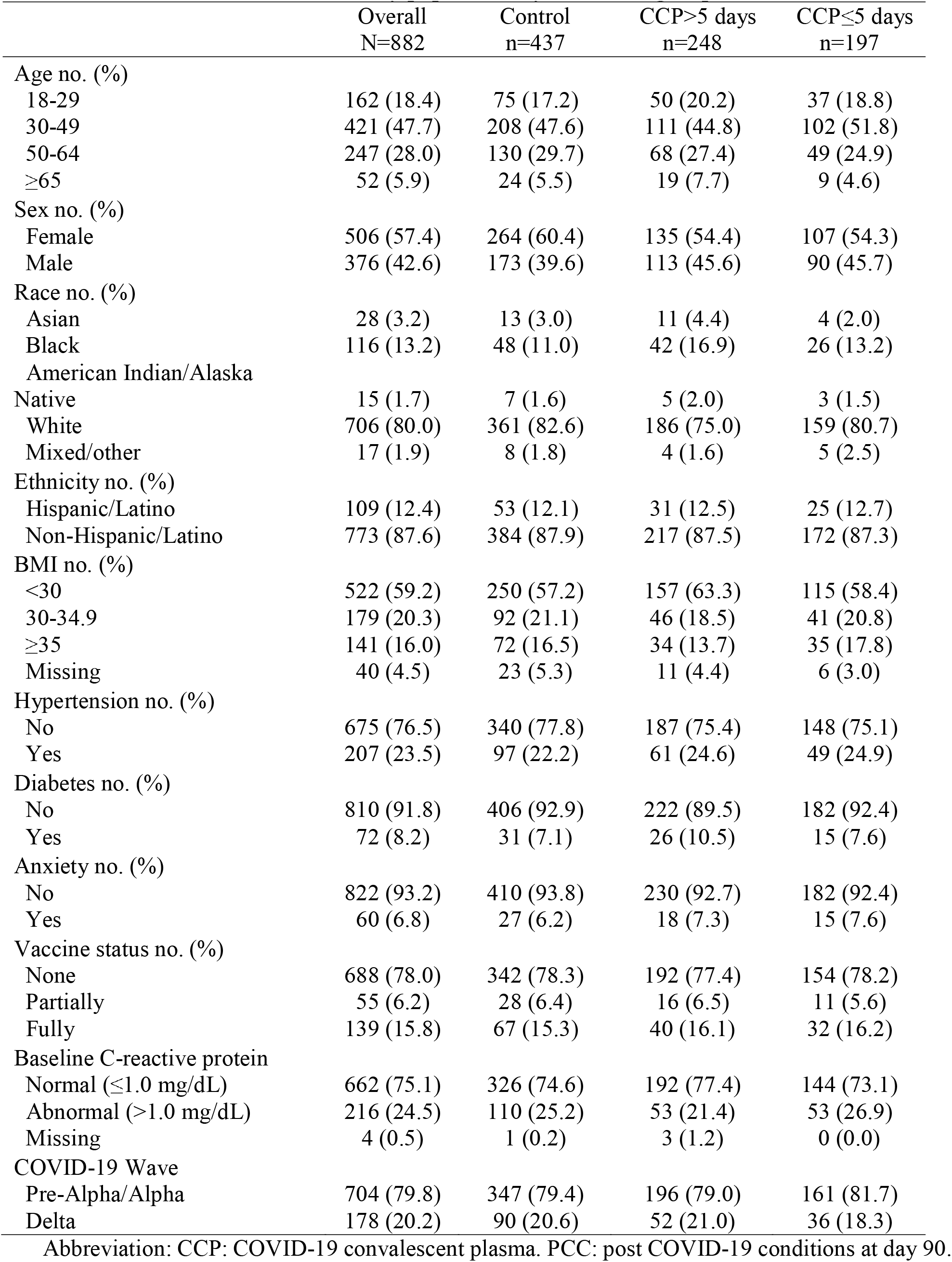
Characteristics of the study population by treatment group.

|  | Overall<br>N=882 | Control<br>n=437 | CCP>5 days<br>n=248 | CCP≤5 days<br>n=197 |
| --- | --- | --- | --- | --- |
| Age no. (%) |  |  |  |  |
| 18-29 | 162 (18.4) | 75 (17.2) | 50 (20.2) | 37 (18.8) |
| 30-49 | 421 (47.7) | 208 (47.6) | 111 (44.8) | 102 (51.8) |
| 50-64 | 247 (28.0) | 130 (29.7) | 68 (27.4) | 49 (24.9) |
| ≥65 | 52 (5.9) | 24 (5.5) | 19 (7.7) | 9 (4.6) |
| Sex no. (%) |  |  |  |  |
| Female | 506 (57.4) | 264 (60.4) | 135 (54.4) | 107 (54.3) |
| Male | 376 (42.6) | 173 (39.6) | 113 (45.6) | 90 (45.7) |
| Race no. (%) |  |  |  |  |
| Asian | 28 (3.2) | 13 (3.0) | 11 (4.4) | 4 (2.0) |
| Black | 116 (13.2) | 48 (11.0) | 42 (16.9) | 26 (13.2) |
| American Indian/Alaska<br>Native | 15 (1.7) | 7 (1.6) | 5 (2.0) | 3 (1.5) |
| White | 706 (80.0) | 361 (82.6) | 186 (75.0) | 159 (80.7) |
| Mixed/other | 17 (1.9) | 8 (1.8) | 4 (1.6) | 5 (2.5) |
| Ethnicity no. (%) |  |  |  |  |
| Hispanic/Latino | 109 (12.4) | 53 (12.1) | 31 (12.5) | 25 (12.7) |
| Non-Hispanic/Latino | 773 (87.6) | 384 (87.9) | 217 (87.5) | 172 (87.3) |
| BMI no. (%) |  |  |  |  |
| <30 | 522 (59.2) | 250 (57.2) | 157 (63.3) | 115 (58.4) |
| 30-34.9 | 179 (20.3) | 92 (21.1) | 46 (18.5) | 41 (20.8) |
| ≥35 | 141 (16.0) | 72 (16.5) | 34 (13.7) | 35 (17.8) |
| Missing | 40 (4.5) | 23 (5.3) | 11 (4.4) | 6 (3.0) |
| Hypertension no. (%) |  |  |  |  |
| No | 675 (76.5) | 340 (77.8) | 187 (75.4) | 148 (75.1) |
| Yes | 207 (23.5) | 97 (22.2) | 61 (24.6) | 49 (24.9) |
| Diabetes no. (%) |  |  |  |  |
| No | 810 (91.8) | 406 (92.9) | 222 (89.5) | 182 (92.4) |
| Yes | 72 (8.2) | 31 (7.1) | 26 (10.5) | 15 (7.6) |
| Anxiety no. (%) |  |  |  |  |
| No | 822 (93.2) | 410 (93.8) | 230 (92.7) | 182 (92.4) |
| Yes | 60 (6.8) | 27 (6.2) | 18 (7.3) | 15 (7.6) |
| Vaccine status no. (%) |  |  |  |  |
| None | 688 (78.0) | 342 (78.3) | 192 (77.4) | 154 (78.2) |
| Partially | 55 (6.2) | 28 (6.4) | 16 (6.5) | 11 (5.6) |
| Fully | 139 (15.8) | 67 (15.3) | 40 (16.1) | 32 (16.2) |
| Baseline C-reactive protein |  |  |  |  |
| Normal (≤1.0 mg/dL) | 662 (75.1) | 326 (74.6) | 192 (77.4) | 144 (73.1) |
| Abnormal (>1.0 mg/dL) | 216 (24.5) | 110 (25.2) | 53 (21.4) | 53 (26.9) |
| Missing | 4 (0.5) | 1 (0.2) | 3 (1.2) | 0 (0.0) |
| COVID-19 Wave |  |  |  |  |
| Pre-Alpha/Alpha | 704 (79.8) | 347 (79.4) | 196 (79.0) | 161 (81.7) |
| Delta | 178 (20.2) | 90 (20.6) | 52 (21.0) | 36 (18.3) |

Abbreviation: CCP: COVID-19 convalescent plasma. PCC: post COVID-19 conditions at day 90.

**Table 2.** Baseline characteristics of the study population by PCC.

|  | Overall<br>N=882 | non-PCC<br>n=590 | PCC<br>n=292 | P value |
| --- | --- | --- | --- | --- |
| Age no. (%) |  |  |  |  |
| 18-29 | 162 (18.4) | 134 (22.7) | 28 (9.6) | **<0.001** |
| 30-49 | 421 (47.7) | 274 (46.4) | 147 (50.3) |  |
| 50-64 | 247 (28.0) | 156 (26.4) | 91 (31.2) |  |
| ≥65 | 52 (5.9) | 26 (4.4) | 26 (8.9) |  |
| Sex no. (%) |  |  |  |  |
| Female | 506 (57.4) | 301 (51.0) | 205 (70.2) | **<0.001** |
| Male | 376 (42.6) | 289 (49.0) | 87 (29.8) |  |
| Race no. (%) |  |  |  |  |
| Asian | 28 (3.2) | 24 (4.1) | 4 (1.4) | 0.061 |
| Black | 116 (13.2) | 74 (12.5) | 42 (14.4) |  |
| American Indian/Alaska Native | 15 (1.7) | 11 (1.9) | 4 (1.4) |  |
| White | 706 (80.0) | 466 (79.0) | 240 (82.2) |  |
| Mixed/other | 17 (1.9) | 15 (2.5) | 2 (0.7) |  |
| Ethnicity no. (%) |  |  |  |  |
| Hispanic/Latino | 109 (12.4) | 66 (11.2) | 43 (14.7) | 0.163 |
| Non-Hispanic/Latino | 773 (87.6) | 524 (88.8) | 249 (85.3) |  |
| BMI no. (%) |  |  |  |  |
| <30 | 522 (59.2) | 365 (61.9) | 157 (53.8) | **0.042** |
| 30-34.9 | 179 (20.3) | 119 (20.2) | 60 (20.5) |  |
| ≥35 | 141 (16.0) | 81 (13.7) | 60 (20.5) |  |
| Missing | 40 (4.5) | 25 (4.2) | 15 (5.1) |  |
| Hypertension no. (%) |  |  |  |  |
| No | 675 (76.5) | 461 (78.1) | 214 (73.3) | 0.130 |
| Yes | 207 (23.5) | 129 (21.9) | 78 (26.7) |  |
| Diabetes no. (%) |  |  |  |  |
| No | 810 (91.8) | 550 (93.2) | 260 (89.0) | **0.045** |
| Yes | 72 (8.2) | 40 (6.8) | 32 (11.0) |  |
| Anxiety no. (%) |  |  |  |  |
| No | 822 (93.2) | 557 (94.4) | 265 (90.8) | 0.059 |
| Yes | 60 (6.8) | 33 (5.6) | 27 (9.2) |  |
| Vaccine status no. (%) |  |  |  |  |
| None | 688 (78.0) | 454 (76.9) | 234 (80.1) | 0.488 |
| Partially | 55 (6.2) | 37 (6.3) | 18 (6.2) |  |
| Fully | 139 (15.8) | 99 (16.8) | 40 (13.7) |  |
| Baseline C-reactive protein |  |  |  |  |
| Normal (≤1.0 mg/dL) | 662 (75.1) | 452 (76.6) | 210 (71.9) | 0.086 |
| Abnormal (>1.0 mg/dL) | 216 (24.5) | 137 (23.2) | 79 (27.1) |  |
| Missing | 4 (0.5) | 1 (0.2) | 3 (1.0) |  |
| Treatment group no. (%) |  |  |  |  |
| Control | 437 (49.5) | 287 (48.6) | 150 (51.4) | 0.149 |
| CCP>5 days since symptom onset | 248 (28.1) | 160 (27.1) | 88 (30.1) |  |
| CCP≤5 days since symptom onset | 197 (22.3) | 143 (24.2) | 54 (18.5) |  |
| COVID-19 Wave |  |  |  |  |
| Pre-Alpha/Alpha | 704 (79.8) | 462 (78.3) | 242 (82.9) | 0.133 |

| Delta | 178 (20.2) | 128 (21.7) | 50 (17.1) |
| --- | --- | --- | --- |

Abbreviation: PCC: Post COVID 19 complication. PCC: post COVID-19 conditions at day 90.
Note: p values for all variables except race were calculated using two-tail $\chi^2$ test, and two-tail fisher's exact test was used for race due to small cell numbers. P values <0.05 were in bold.

### SOLUBLE MARKERS OF INFLAMMATION

Levels of most cytokines decreased over time from baseline screening to day 90 (**Figure 2, Supplemental Figure 1**). Of the 21 cytokines evaluated, IL-1RA, IL-6, IL-8, IL-15 and MCP-1 were elevated at baseline among those with PCC compared to participants who did not develop PCC (**Figure 3, Supplemental Figure 2**). Elevated levels of IL-6 were significantly associated with PCC after correcting for multiple comparisons. In a multivariable model adjusting for age, sex, race, BMI, diabetes, vaccine status, and plasma transfused, an elevated level of IL-6 at baseline was associated with the presence of PCC (Adjusted Odds Ratio [AOR]=1.59, 95%CI=1.02-2.47) (**Supplemental Table 5)**.

**Figure 2.**
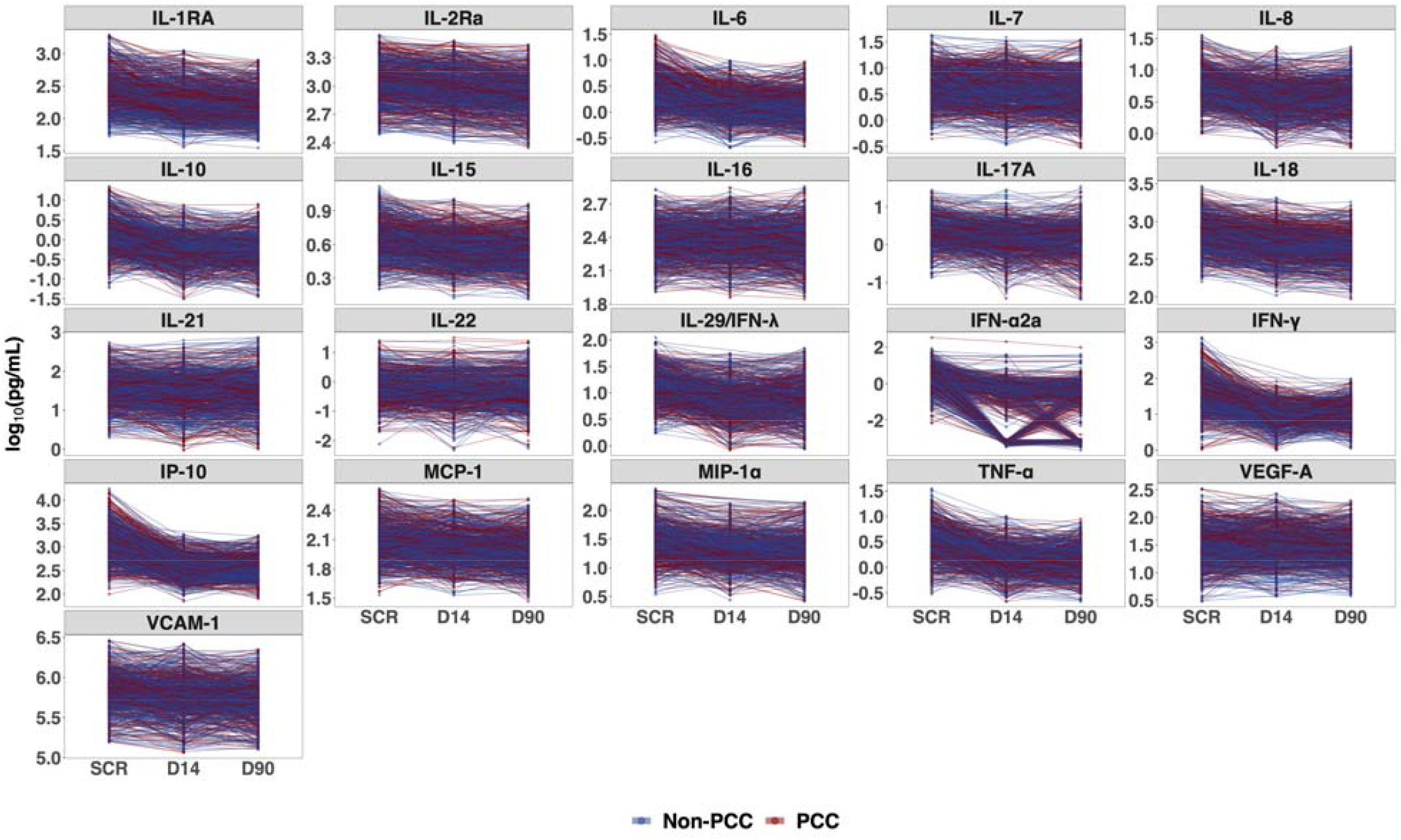
Trajectory of the cytokines during study period. Abbreviation: PCC: Post COVID 19 condition. SCR: screening visit. D14: Day 14 visit. D90: day 90 visit. Note: Each dot represents a sample, and each line represents a person.

**Figure 3.**
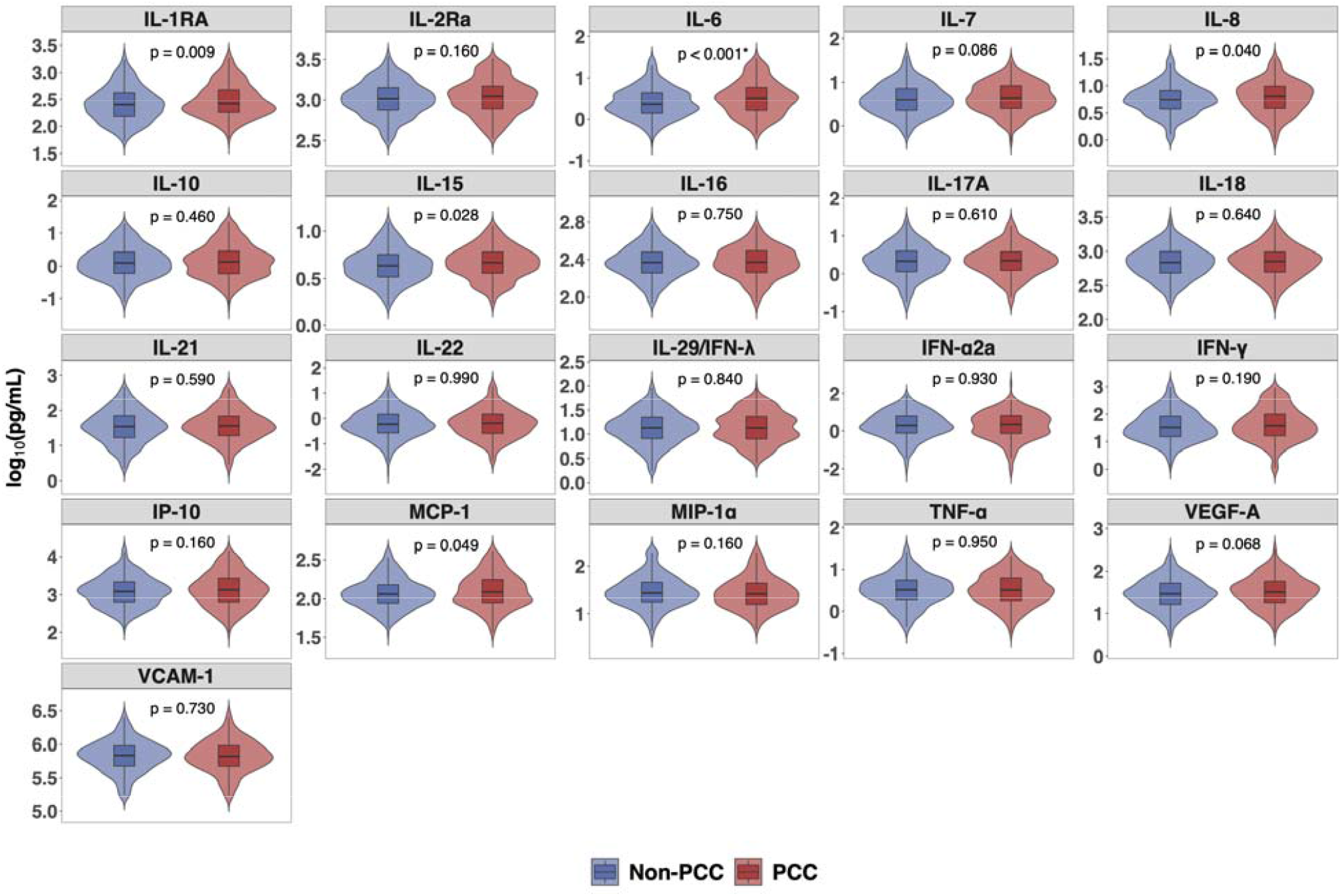
Comparison of cytokines at screening between study participants with PCC and without PCC. Abbreviation: PCC: Post COVID-19 condition. Note: P values were determined by Wilcoxon rank sum test. *IL-6 were significantly different after adjusting multiple comparison using Benjamini-Hockberg correction with a false discovery rate of 0.05.

### BASELINE FACTORS ASSOCIATED WITH PCC

In multivariable analysis, (n=882) older age, female sex, and baseline elevated IL-6 were associated with PCC. (**Table 3a**). Early treatment with CCP (≤5 days from symptom onset) trended towards a lower odds of PCC (AOR=0.73 [0.48, 1.11]) compared to those who received control plasma, When evaluating only those who received CCP, those who received early treatment had statistically significant lower odds of PCC (AOR=0.60 [0.38, 0.95]) compared to those who received late CCP treatment (**Table 3b**). There was no statistically significant interaction between CCP treatment and IL-6. Similar trends, although not significant, were seen among the full trial population seen at day 90 (N=1061) (**Supplemental Tables 6a & 6b**).

**Table 3a.** Logistic regression model assessing demographic and clinical factors and PCC among the study population.

| Analyte | OR (95% CI), n = 882 |  |  |
| --- | --- | --- | --- |
|  | Univariate | Model 1 | Model 2 |
| Age, scaled to 10 years | **1.31 (1.18, 1.46)** | **1.35 (1.20, 1.51)** | **1.32 (1.17, 1.50)** |
| Sex |  |  |  |
| Male | Ref. | Ref. | Ref. |
| Female | **2.26 (1.68, 3.06)** | **2.38 (1.74, 3.28)** | **2.70 (1.93, 3.81)** |
| Race |  |  |  |
| White | Ref. | Ref. | Ref. |
| Non-white | 0.84 (0.58, 1.20) | 0.81 (0.54, 1.19) | 0.85 (0.56, 1.28) |
| BMI |  |  |  |
| <30 | Ref. | Ref. | Ref. |
| ≥30 | **1.39 (1.04, 1.87)** | 1.20 (0.88, 1.65) | 1.19 (0.85, 1.67) |
| Vaccine Status |  |  |  |
| None | Ref. | Ref. | Ref. |
| Partially or fully | 0.83 (0.58, 1.16) | 0.87 (0.60, 1.25) | 0.86 (0.59, 1.26) |
| Diabetes |  |  |  |
| No | Ref. | Ref. | Ref. |
| Yes | **1.69 (1.03, 2.75)** | 1.25 (0.72, 2.15) | 1.24 (0.68, 2.21) |
| Plasma transfused |  |  |  |
| Control plasma | Ref. | Ref. | Ref. |
| CCP>5 days since symptom onset | 1.05 (0.76, 1.46) | 1.24 (0.87, 1.76) | 1.18 (0.81, 1.72) |
| CCP≤5 days since symptom onset | 0.72 (0.50, 1.04) | 0.81 (0.54, 1.20) | 0.73 (0.48, 1.11) |
| Log<sub>10</sub> IL-6, pg/mL | - | - | **1.59 (1.02, 2.47)** |

**Table 3b.**
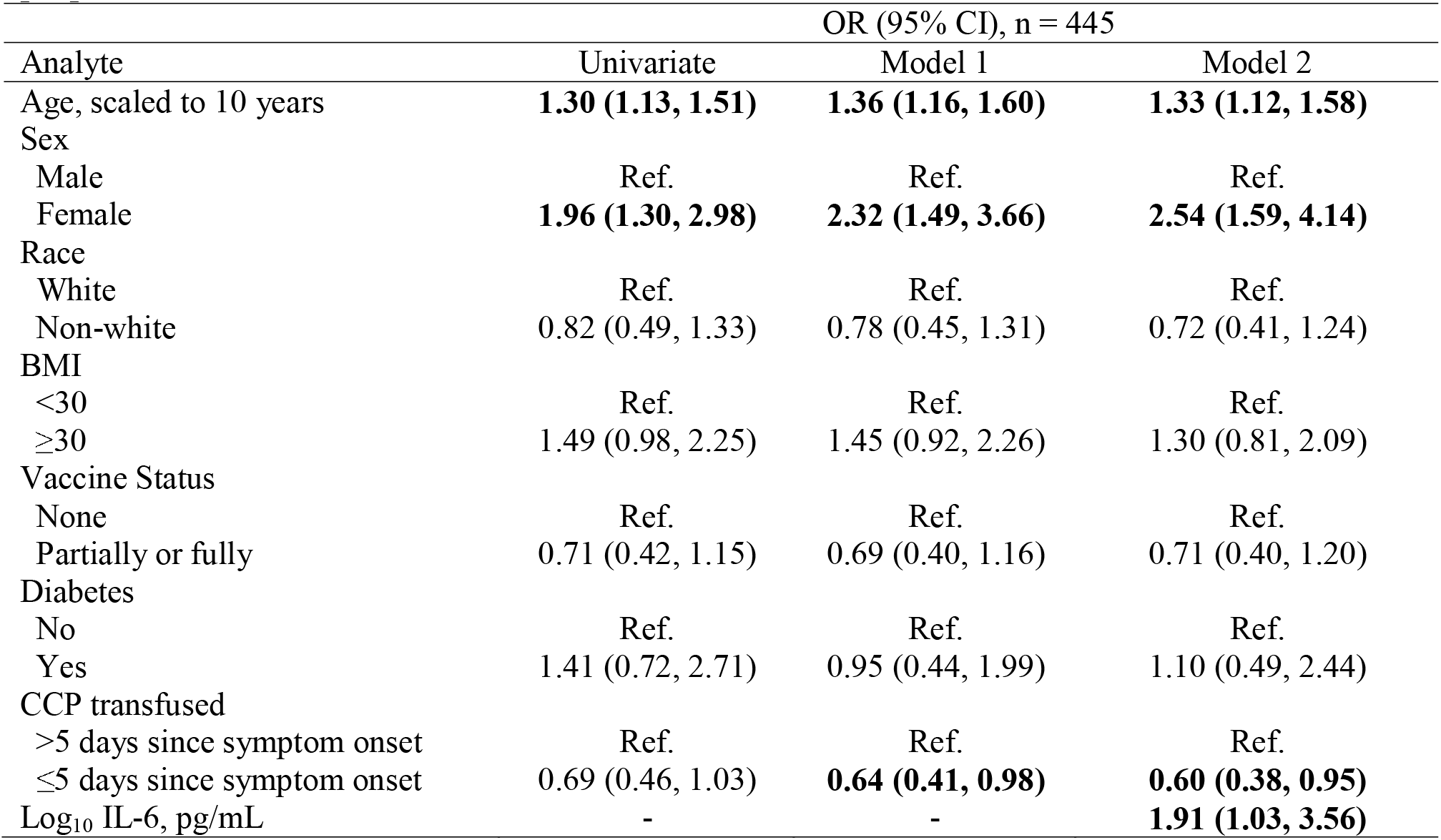
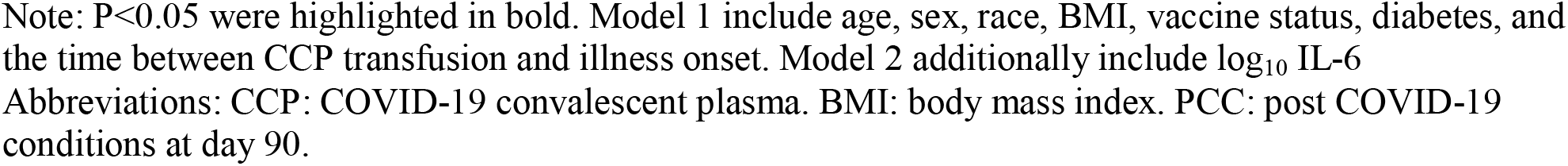
Logistic regression model assessing demographic and clinical factors and PCC among the people transfused with CCP.

| Analyte | OR (95% CI), n = 445 |  |  |
| --- | --- | --- | --- |
|  | Univariate | Model 1 | Model 2 |
| Age, scaled to 10 years | **1.30 (1.13, 1.51)** | **1.36 (1.16, 1.60)** | **1.33 (1.12, 1.58)** |
| Sex |  |  |  |
| Male | Ref. | Ref. | Ref. |
| Female | **1.96 (1.30, 2.98)** | **2.32 (1.49, 3.66)** | **2.54 (1.59, 4.14)** |
| Race |  |  |  |
| White | Ref. | Ref. | Ref. |
| Non-white | 0.82 (0.49, 1.33) | 0.78 (0.45, 1.31) | 0.72 (0.41, 1.24) |
| BMI |  |  |  |
| <30 | Ref. | Ref. | Ref. |
| ≥30 | 1.49 (0.98, 2.25) | 1.45 (0.92, 2.26) | 1.30 (0.81, 2.09) |
| Vaccine Status |  |  |  |
| None | Ref. | Ref. | Ref. |
| Partially or fully | 0.71 (0.42, 1.15) | 0.69 (0.40, 1.16) | 0.71 (0.40, 1.20) |
| Diabetes |  |  |  |
| No | Ref. | Ref. | Ref. |
| Yes | 1.41 (0.72, 2.71) | 0.95 (0.44, 1.99) | 1.10 (0.49, 2.44) |
| CCP transfused |  |  |  |
| >5 days since symptom onset | Ref. | Ref. | Ref. |
| ≤5 days since symptom onset | 0.69 (0.46, 1.03) | **0.64 (0.41, 0.98)** | **0.60 (0.38, 0.95)** |
| Log<sub>10</sub> IL-6, pg/mL | - | - | **1.91 (1.03, 3.56)** |

Note: $P < 0.05$ were highlighted in bold. Model 1 include age, sex, race, BMI, vaccine status, diabetes, and the time between CCP transfusion and illness onset. Model 2 additionally include $\log_{10}$ IL-6
Abbreviations: CCP: COVID-19 convalescent plasma. BMI: body mass index. PCC: post COVID-19 conditions at day 90.

## DISCUSSION

This study is among the first to show elevation of IL-6 at infection was associated with PCC and that early treatment with CCP for COVID-19 trended towards a lower odds of PCC. Notably, greater levels of IL-6 at baseline were associated with the presence of symptoms at day 90, while early treatment with CCP was associated with lower odds of PCC. Finally, consistent with other studies, advanced age [10], diabetes mellitus, and higher BMI were associated with PCC [6, 11, 31].

Approximately one-third of participants with outpatient SARS-CoV-2 infection diagnosed early in the pandemic continued to have symptoms 90 days after infection. The most common symptoms at day 90 were less likely to be respiratory, and more consistent with neurologic injury including fatigue, loss of smell, and loss of taste [12]. A recent Veterans Affairs prepublication study demonstrated that treatment with nirmatrelvir during the acute phase reduces the risk of PCC [13]. CCP has been shown to reduce hospitalizations and prevent morbidity and mortality when provided early and at high-titer [32-39]. In this early treatment trial, CCP reduced COVID-19 related hospitalizations by 54% and demonstrated a trend towards lower odds of PCC overall, and even lower PCC odds among those treated with CCP within 5 days [27]. With larger sample size, it may be possible to identify patient populations or clinical phenotypes of PCC that may benefit from early treatment to prevent PCC. Further understanding is required of how acute SARS-CoV-2 infection injures organs, the implications of viral persistence of SARS-CoV-2 in tissues, and the immune dysregulation that can occur after COVID-19. Additional studies are needed to better understand the impact of early treatment with monoclonals, CCP, remdesivir, and other oral antivirals among those with outpatient SARS-CoV-2 infection.

IL-6 plays a critical role during infections; this proinflammatory cytokine leads to an acute-phase response, B-cell maturation and also T-cell expansion [40]. Notably, elevated levels of IL-6 at screening were associated with the development of PCC after adjusting for demographic factors and comorbidities. While cytokine levels decreased from baseline screening to day 90, they remained elevated during the COVID-19 recovery phase, which likely contributes to PCC.

This study had several notable strengths. First, the study population was large, geographically and demographically diverse, and entirely outpatient [27]. Second, biospecimens and data on symptoms were actively ascertained in real time from participants through in-person visits early in the onset of illness and on day 90 after transfusion. In addition to the presence or absence of symptoms over time, we collected samples at each visit to allow for cytokine analysis. Finally, study data on symptoms were collected uniformly across all sites, including pre-omicron COVID-19 variants, thus increasing the generalizability of our findings.

This study has several important limitations. First, participants were asked about seventeen symptoms identified early in the trial as important symptoms of COVID-19. During the pandemic, knowledge of PCC expanded to include more neurocognitive symptoms; however, we could not conduct additional validated measurements of sleep or neurologic function as many participants had completed follow-up. Also, while 90-day follow-up was excellent (90%), it was not complete. However, those who did not complete the study did not differ demographically from those who did. In addition, COVID-19 has disproportionately affected the elderly and people of color. Within our sample, only a small proportion were age ≥ 65 years or of minority ethnicity or race, though the proportions were similar to those in the general American population. Our study was also limited by sample size in trying to understand the impact of early treatment on the various clinical phenotypes associated with PCC. We could not further delineate PCC phenotype beyond the presence or absence of symptoms at a fixed time. More inclusive phenotyping including exacerbation of a chronic condition or development of a new comorbid condition during recovery may yield different results. Also, this study was done early in the pandemic when relatively few participants were fully vaccinated. Results may differ among participants who have immunity from prior infection or vaccination.

In summary, this study demonstrated high rates of PCC symptoms at day 90, particularly among those with higher baseline levels of IL-6 at the time of infection or who did not receive early CCP treatment. Future studies might examine the impact of anti-IL6 agents and the development of PCC among outpatients. Despite increasing vaccination uptake, PCC risk does not disappear, and IL-6 modulation may be a possible therapeutic intervention to reduce the burden of long-term symptoms among those with SARS-CoV-2 infection.

## Data Availability

All data produced in the present work are contained in the manuscript

## Acknowledgments

The authors gratefully acknowledge the plasma donors and study participants who generously gave of their time and biological specimens to the CSSC-001 and CSSC-004 trials and the passionate study personnel who facilitated these studies.

## Disclaimer

The content is solely the responsibility of the authors and does not necessarily represent the official views of the funders.

## Conflicts of Interest

**Kelly Gebo**- Consults for the Aspen Institute, Teach for America, served as a non-paid member of scientific advisory board for Pfizer and writes COVID management guidelines for UpToDate which are out of scope of paper.

**Sonya L. Heath**- Nothing to disclose

**Yuriko Fukuta**- Nothing to disclose

**Xianming Zhu**- Nothing to disclose

**Sheriza Baksh**- Nothing to disclose

**Alison G. Abraham**- Consultant for Implementation Group Inc, Hirslanden Klinik, Zurich CH, & ELSEVIER,

**Feben Habtehyimer**- Nothing to disclose

**David Shade**- Nothing to disclose

**Jessica Ruff**- Nothing to disclose

**Malathi Ram**- Nothing to disclose

**Oliver Laeyendecker**- Nothing to disclose

**Reinaldo E. Fernandez**- Nothing to disclose

**Eshan U. Patel**- Nothing to disclose

**Owen R. Baker**- Nothing to disclose

**Shmuel Shoham**- Served on a CCP guideline panel

**Edward R. Cachay**- has received unrestricted research grants from Gilead and Merck paid to the Regents of the University of California. He also participated in an advisory board to Theratechnologies for an unrelated topic.

**Judith S. Currier**- Consulted for Merck and Company in 2021, currently not on any guidelines panel

**Jonathan M. Gerber**- Nothing to disclose

**Thomas J. Gniadek**- Currently employed by Fenwal, Inc., a Fresenius Kabi Company.

**Barry Meisenberg**- Nothing to disclose

**Donald N. Forthal**- nothing to disclose

**Laura L. Hammitt-** research funding to my institution from AstraZeneca, CDC, Merck, NIH, and Pfizer.

**Moises A. Huaman**- M.A.H. reports contracts from Gilead Sciences Inc, Insmed Inc, AN2 Therapeutics, Inc to the University of Cincinnati, outside the submitted work.

**Adam Levine**- Nothing to disclose

**Giselle S. Mosnaim**- Received research grant support from Teva, Alk-Abello, and Genentech and currently receives research grant support from Novartis, GlaxoSmithKline, and Sanofi-Regeneron. Serves as Immediate Past President of the American Academy of Allergy Asthma and Immunology and Co-Chair of the Continuous Assessment Program Examination for the American Board of Allergy and Immunology

**Bela Patel**- Part of COVID trials and PAH trials, no disclosures relevant to CP

**James H. Paxton**- Research funding from MindRhythm, Inc

**Jay S. Raval**- Consultant and Advisor with Sanofi Genzyme; Board of Directors Member with the American Society for Apheresis, no overlap with CP

**Catherine G. Sutcliffe**- Nothing to disclose

**Shweta Anjan**- Nothing to disclose

**Seble Kassaye**- Helped to produce educational materials related to HIV with Integritas Communications, LLC and Vindico Medical Education, LLC

**Janis E. Blair**- Nothing to disclose

**Karen Lane**- nothing to disclose

**Nichol A. McBee**- Nothing to disclose

**Amy L. Gawad**- Nothing to disclose

**Piyali Das**- Nothing to disclose

**Sabra L. Klein**- Nothing to disclose

**Andrew Pekosz**- Nothing to disclose

**Arturo Casadevall**- Serve on the scientific advisory board of SAB Therapeutics

**Evan M. Bloch**- EMB reports personal fees and non-financial support from Terumo BCT, Abbott Laboratories, Tegus and UptoDate, outside of the submitted work. EMB is a member of the United States Food and Drug Administration (FDA) Blood Products Advisory Committee. Served on a CCP guideline panel

**Daniel Hanley**- Dr. Hanley reports personal fees from Neurelis, Neurotrope, and medicolegal consulting.

**Aaron A.R. Tobian**- Served on a CCP guideline panel

**David J. Sullivan**- Founder and Board member with stock options (macrolide for malaria) DJS reports AliquantumRx, Hemex Health malaria diagnostics consulting and royalties for malaria diagnostic test control standards to Alere-all outside of submitted work

