## Supplemental Tables and FIgure for "Early Treatment, Inflammation and Post-COVID Conditions"

This appendix has been provided by the authors to give readers additional information.

Table of Contents	1

Enrolling Centers, Principal Investigators	2

Supplement Table 1	Supplemental Table 1. Comparison of the baseline characteristics between the study population and the excluded population	4

Supplement Table 2	Characteristics of the study population by treatment group among those have day 90 symptom data	6

Supplement Table 3	Characteristics of the study population by treatment group among the

Trial	8

Supplement Table 4	Symptoms at day 90 visit among the study population	10

Supplement Figure 1	Trajectory of the cytokines during study period (outliers included)	11

Supplemental Figure 2 Comparison of cytokines between PCC and non-PCC at screening, day 14 and day 90	12

Supplemental Table 5 Multivariable logistic regression between log_10_ cytokine levels at screening and PCC	13

Supplemental Table 6a. Logistic regression between demographic and clinical factors and PCC among the people in the original trial and have day 90 symptom data	14

Supplemental Table 6b. Logistic regression between demographic and clinical factors and PCC among the people having day 90 symptom data and transfused with CCP in the original trial 15

The members of the COVID-10 Serologic Studies Consortium (CSSC) are: Anne Arundel Medical Center (Barry R. Meisenberg); Ascada Research (Matthew Abinante, Kevin Oei); Baylor College of Medicine (Yuriko Fukuta); MedStar Georgetown University Hospital (Seble Kessaye); Johns Hopkins Center for American Indian Health (Laura L. Hammitt, Catherine G. Sutcliffe) Johns Hopkins Bloomberg School of Public Health (David J Sullivan, Bryan Lau, Atika Singh, David M. Shade, Stephan Ehrhardt, Sheriza N. Baksh, Andrew Pekosz, Sabra L. Klein, Arturo Casadevall); Johns Hopkins University (Kelly A Gebo, Shmuel Shoham, Evan M. Bloch, Christi E. Marshall, Anusha Yarava, Karen Lane, Nichol A. McBee, Amy L. Gawad, Nicky Karlen, Daniel E. Ford, Douglas A. Jabs, Lawrence J. Appel, Oliver Laeyendecker, Aaron A.R. Tobian, Daniel F. Hanley); Lifespan/Brown University Rhode Island Hospital (Adam C. Levine); Mayo Clinic, Phoenix (Janis E. Blair); MedStar Washington Hospital Center (Aarthi Shenoy); NorthShore University HealthSystem (Giselle S. Mosnaim, Thomas J. Gniadek); The Bliss Group (Michael Roth); The Next Practice Group (Colin Foster); University of California Los Angeles (Judith S. Currier); University of Alabama at Birmingham (Sonya L. Health); University of California, Irvine Health (Donald N. Forthal); University of California, San Diego (Edward R. Cachay, Elizabeth S. Allen); University of Cincinnati Medical Center (Moises A. Huaman); University of Massachusetts Worcester (Jonathan M. Gerber); University of Miami (Shweta Anjan); University of New Mexico (Jay S. Raval); University of Rochester (Martin S. Zand); University of Texas Health Science Center at Houston (Bela Patel); University of Utah Health (Emily S. Spivak, J. Robinson Singleton); Vassar Brothers Medical Center (Valerie C. Cluzet, Daniel Cruser); Wayne State University (James H. Paxton); Western Connecticut Health Network, Danbury Hospital (Joann R. Petrini, Patrick B. Broderick, William Rausch, MarieElena Cordisco); Western Connecticut Health Network, Norwalk Hospital (Jean Hammel, Benjamin Greenblatt)

**Supplemental Table 1. Comparison of the baseline characteristics between the study population and the excluded population.**

|  | Overall  N=1181 | Cytokine tested,  n=882 | Cytokine not tested,  n=299 |
| --- | --- | --- | --- |
| Age no. % |  |  |  |
| 18-29 | 218 (18.5) | 162 (18.4) | 56 (18.7) |
| 30-49 | 552 (46.7) | 421 (47.7) | 131 (43.8) |
| 50-64 | 331 (28.0) | 247 (28.0) | 84 (28.1) |
| ≥65 | 80 (6.8) | 52 (5.9) | 28 (9.4) |
| Sex no. % |  |  |  |
| Female | 675 (57.2) | 506 (57.4) | 169 (56.5) |
| Male | 506 (42.8) | 376 (42.6) | 130 (43.5) |
| Race no. % |  |  |  |
| Asian | 44 (3.7) | 28 (3.2) | 16 (5.4) |
| Black | 162 (13.7) | 116 (13.2) | 46 (15.4) |
| Mixed/other | 25 (2.1) | 17 (1.9) | 8 (2.7) |
| American Indian/Alaska Native | 17 (1.4) | 15 (1.7) | 2 (0.7) |
| White | 933 (79.0) | 706 (80.0) | 227 (75.9) |
| Ethnicity no. % |  |  |  |
| Hispanic/Latino | 170 (14.4) | 109 (12.4) | 61 (20.4) |
| Non-Hispanic/Latino | 1011 (85.6) | 773 (87.6) | 238 (79.6) |
| BMI no. % |  |  |  |
| <30 | 679 (57.5) | 522 (59.2) | 157 (52.5) |
| 30-34.9 | 240 (20.3) | 179 (20.3) | 61 (20.4) |
| ≥35 | 204 (17.3) | 141 (16.0) | 63 (21.1) |
| Missing | 58 (4.9) | 40 (4.5) | 18 (6.0) |
| Hypertension no. % |  |  |  |
| No | 905 (76.6) | 675 (76.5) | 230 (76.9) |
| Yes | 276 (23.4) | 207 (23.5) | 69 (23.1) |
| Diabetes no. % |  |  |  |
| No | 1082 (91.6) | 810 (91.8) | 272 (91.0) |
| Yes | 99 (8.4) | 72 (8.2) | 27 (9.0) |
| Anxiety no. % |  |  |  |
| No | 1106 (93.6) | 822 (93.2) | 284 (95.0) |
| Yes | 75 (6.4) | 60 (6.8) | 15 (5.0) |
| Vaccine status no. % |  |  |  |
| None | 964 (81.6) | 688 (78.0) | 276 (92.3) |
| Partially | 58 (4.9) | 55 (6.2) | 3 (1.0) |
| Fully | 159 (13.5) | 139 (15.8) | 20 (6.7) |
| C-reactive protein |  |  |  |
| Normal (≤1.0 mg/dL) | 861 (72.9) | 662 (75.1) | 199 (66.6) |
| Abnormal (>1.0 mg/dL) | 309 (26.2) | 216 (24.5) | 93 (31.1) |
| Missing | 11 (0.9) | 4 (0.5) | 7 (2.3) |
| Treatment group no. % |  |  |  |
| Control | 589 (49.9) | 437 (49.5) | 152 (50.8) |
| CCP>5 days since symptom onset | 335 (28.4) | 248 (28.1) | 87 (29.1) |
| CCP≤5 days since symptom onset | 257 (21.8) | 197 (22.3) | 60 (20.1) |
| COVID-19 Wave |  |  |  |
| Pre-Alpha/Alpha | 947 (80.2) | 704 (79.8) | 243 (81.3) |
| Delta | 234 (19.8) | 178 (20.2) | 56 (18.7) |

Abbreviation: CCP: COVID-19 convalescent plasma. PCC: post COVID-19 conditions at day 90.

**Supplemental Table 2. Characteristics of the study population by treatment group among those have day 90 symptom data**

|  | Overall  N=1061 | Control  n=528 | CCP>5 days  n=301 | CCP≤5 days  n=232 |
| --- | --- | --- | --- | --- |
| Age |  |  |  |  |
| 18-29 | 183 (17.2) | 83 (15.7) | 57 (18.9) | 43 (18.5) |
| 30-49 | 491 (46.3) | 245 (46.4) | 129 (42.9) | 117 (50.4) |
| 50-64 | 313 (29.5) | 165 (31.2) | 88 (29.2) | 60 (25.9) |
| ≥65 | 74 (7.0) | 35 (6.6) | 27 (9.0) | 12 (5.2) |
| Sex |  |  |  |  |
| Female | 607 (57.2) | 316 (59.8) | 163 (54.2) | 128 (55.2) |
| Male | 454 (42.8) | 212 (40.2) | 138 (45.8) | 104 (44.8) |
| Race |  |  |  |  |
| Asian | 35 (3.3) | 16 (3.0) | 14 (4.7) | 5 (2.2) |
| Black | 142 (13.4) | 62 (11.7) | 50 (16.6) | 30 (12.9) |
| American Indian/Alaska Native | 17 (1.6) | 9 (1.7) | 5 (1.7) | 3 (1.3) |
| White | 846 (79.7) | 429 (81.2) | 228 (75.7) | 189 (81.5) |
| Mixed/other | 21 (2.0) | 12 (2.3) | 4 (1.3) | 5 (2.2) |
| Ethnicity |  |  |  |  |
| Hispanic/Latino | 134 (12.6) | 71 (13.4) | 34 (11.3) | 29 (12.5) |
| Non-Hispanic/Latino | 927 (87.4) | 457 (86.6) | 267 (88.7) | 203 (87.5) |
| BMI |  |  |  |  |
| <30 | 616 (58.1) | 294 (55.7) | 184 (61.1) | 138 (59.5) |
| 30-34.9 | 213 (20.1) | 114 (21.6) | 56 (18.6) | 43 (18.5) |
| ≥35 | 178 (16.8) | 94 (17.8) | 42 (14.0) | 42 (18.1) |
| Missing | 54 (5.1) | 26 (4.9) | 19 (6.3) | 9 (3.9) |
| Hypertension |  |  |  |  |
| No | 799 (75.3) | 401 (75.9) | 227 (75.4) | 171 (73.7) |
| Yes | 262 (24.7) | 127 (24.1) | 74 (24.6) | 61 (26.3) |
| Diabetes |  |  |  |  |
| No | 968 (91.2) | 483 (91.5) | 272 (90.4) | 213 (91.8) |
| Yes | 93 (8.8) | 45 (8.5) | 29 (9.6) | 19 (8.2) |
| Anxiety |  |  |  |  |
| No | 991 (93.4) | 495 (93.8) | 281 (93.4) | 215 (92.7) |
| Yes | 70 (6.6) | 33 (6.2) | 20 (6.6) | 17 (7.3) |
| Vaccine status |  |  |  |  |
| None | 856 (80.7) | 426 (80.7) | 244 (81.1) | 186 (80.2) |
| Partially | 56 (5.3) | 29 (5.5) | 16 (5.3) | 11 (4.7) |
| Fully | 149 (14.0) | 73 (13.8) | 41 (13.6) | 35 (15.1) |
| Baseline C-reactive protein |  |  |  |  |
| Normal (≤1.0 mg/dL) | 779 (73.4) | 390 (73.9) | 223 (74.1) | 166 (71.6) |
| Abnormal (>1.0 mg/dL) | 275 (25.9) | 137 (25.9) | 74 (24.6) | 64 (27.6) |
| Missing | 7 (0.7) | 1 (0.2) | 4 (1.3) | 2 (0.9) |
| COVID-19 Wave |  |  |  |  |
| Pre-Alpha/Alpha | 867 (81.7) | 429 (81.2) | 245 (81.4) | 193 (83.2) |
| Delta | 194 (18.3) | 99 (18.8) | 56 (18.6) | 39 (16.8) |

Abbreviation: CCP: COVID-19 convalescent plasma. PCC: post COVID-19 conditions at day 90.

**Supplemental Table 3. Characteristics of the study population by treatment group among the trial**

|  | Overall  N=1181 | Control  n=589 | CCP>5 days  n=335 | CCP≤5 days  n=257 |
| --- | --- | --- | --- | --- |
| Age no. % |  |  |  |  |
| 18-29 | 218 (18.5) | 98 (16.6) | 65 (19.4) | 55 (21.4) |
| 30-49 | 552 (46.7) | 275 (46.7) | 151 (45.1) | 126 (49.0) |
| 50-64 | 331 (28.0) | 176 (29.9) | 91 (27.2) | 64 (24.9) |
| ≥65 | 80 (6.8) | 40 (6.8) | 28 (8.4) | 12 (4.7) |
| Sex no. % |  |  |  |  |
| Female | 675 (57.2) | 352 (59.8) | 182 (54.3) | 141 (54.9) |
| Male | 506 (42.8) | 237 (40.2) | 153 (45.7) | 116 (45.1) |
| Race no. % |  |  |  |  |
| Asian | 44 (3.7) | 22 (3.7) | 15 (4.5) | 7 (2.7) |
| Black | 162 (13.7) | 70 (11.9) | 58 (17.3) | 34 (13.2) |
| American Indian/Alaska Native | 17 (1.4) | 9 (1.5) | 5 (1.5) | 3 (1.2) |
| White | 933 (79.0) | 475 (80.6) | 252 (75.2) | 206 (80.2) |
| Mixed/other | 25 (2.1) | 13 (2.2) | 5 (1.5) | 7 (2.7) |
| Ethnicity no. % |  |  |  |  |
| Hispanic/Latino | 170 (14.4) | 90 (15.3) | 43 (12.8) | 37 (14.4) |
| Non-Hispanic/Latino | 1011 (85.6) | 499 (84.7) | 292 (87.2) | 220 (85.6) |
| BMI no. % |  |  |  |  |
| <30 | 679 (57.5) | 325 (55.2) | 200 (59.7) | 154 (59.9) |
| 30-34.9 | 240 (20.3) | 127 (21.6) | 66 (19.7) | 47 (18.3) |
| ≥35 | 204 (17.3) | 107 (18.2) | 50 (14.9) | 47 (18.3) |
| Missing | 58 (4.9) | 30 (5.1) | 19 (5.7) | 9 (3.5) |
| Hypertension |  |  |  |  |
| No | 905 (76.6) | 453 (76.9) | 257 (76.7) | 195 (75.9) |
| Yes | 276 (23.4) | 136 (23.1) | 78 (23.3) | 62 (24.1) |
| Diabetes |  |  |  |  |
| No | 1082 (91.6) | 539 (91.5) | 305 (91.0) | 238 (92.6) |
| Yes | 99 (8.4) | 50 (8.5) | 30 (9.0) | 19 (7.4) |
| Anxiety |  |  |  |  |
| No | 1106 (93.6) | 554 (94.1) | 313 (93.4) | 239 (93.0) |
| Yes | 75 (6.4) | 35 (5.9) | 22 (6.6) | 18 (7.0) |
| Vaccine status |  |  |  |  |
| None | 964 (81.6) | 479 (81.3) | 275 (82.1) | 210 (81.7) |
| Partially | 58 (4.9) | 31 (5.3) | 16 (4.8) | 11 (4.3) |
| Fully | 159 (13.5) | 79 (13.4) | 44 (13.1) | 36 (14.0) |
| Baseline C-reactive protein |  |  |  |  |
| Normal (≤1.0 mg/dL) | 861 (72.9) | 432 (73.3) | 247 (73.7) | 182 (70.8) |
| Abnormal (>1.0 mg/dL) | 309 (26.2) | 155 (26.3) | 82 (24.5) | 72 (28.0) |
| Missing | 11 (0.9) | 2 (0.3) | 6 (1.8) | 3 (1.2) |
| COVID-19 Wave |  |  |  |  |
| Pre-Alpha/Alpha | 947 (80.2) | 473 (80.3) | 266 (79.4) | 208 (80.9) |
| Delta | 234 (19.8) | 116 (19.7) | 69 (20.6) | 49 (19.1) |

Abbreviation: CCP: COVID-19 convalescent plasma. PCC: post COVID-19 conditions at day 90.

**Supplemental table 4. Symptoms at day 90 visit among the study population**

| Symptoms, no. % | N=882 (%) |
| --- | --- |
| Anosmia | 128 (14.5) |
| Ageusia | 88 (10.0) |
| Chills | 4 (0.5) |
| Cough | 63 (7.1) |
| Diarrhea | 13 (1.5) |
| Fatigue | 128 (14.5) |
| Fever | 0 (0.0) |
| Headache | 59 (6.7) |
| Myalgia | 39 (4.4) |
| Nausea | 21 (2.4) |
| Neurological complication | 79 (9.0) |
| Runny nose | 34 (3.9) |
| Shortness of breath | 70 (7.9) |
| Skin manifestations | 10 (1.1) |
| Sore throat | 11 (1.2) |
| Stuffy nose | 36 (4.1) |
| Vomiting | 1 (0.1) |

**Supplemental figure 1. Trajectory of the cytokines during study period (outliers included)**

.

Abbreviation: PCC: Post COVID 19 condition. SCR: screening visit. D14: Day 14 visit. D90: day 90 visit.

Note: Each dot represents a sample, and each line represents a person
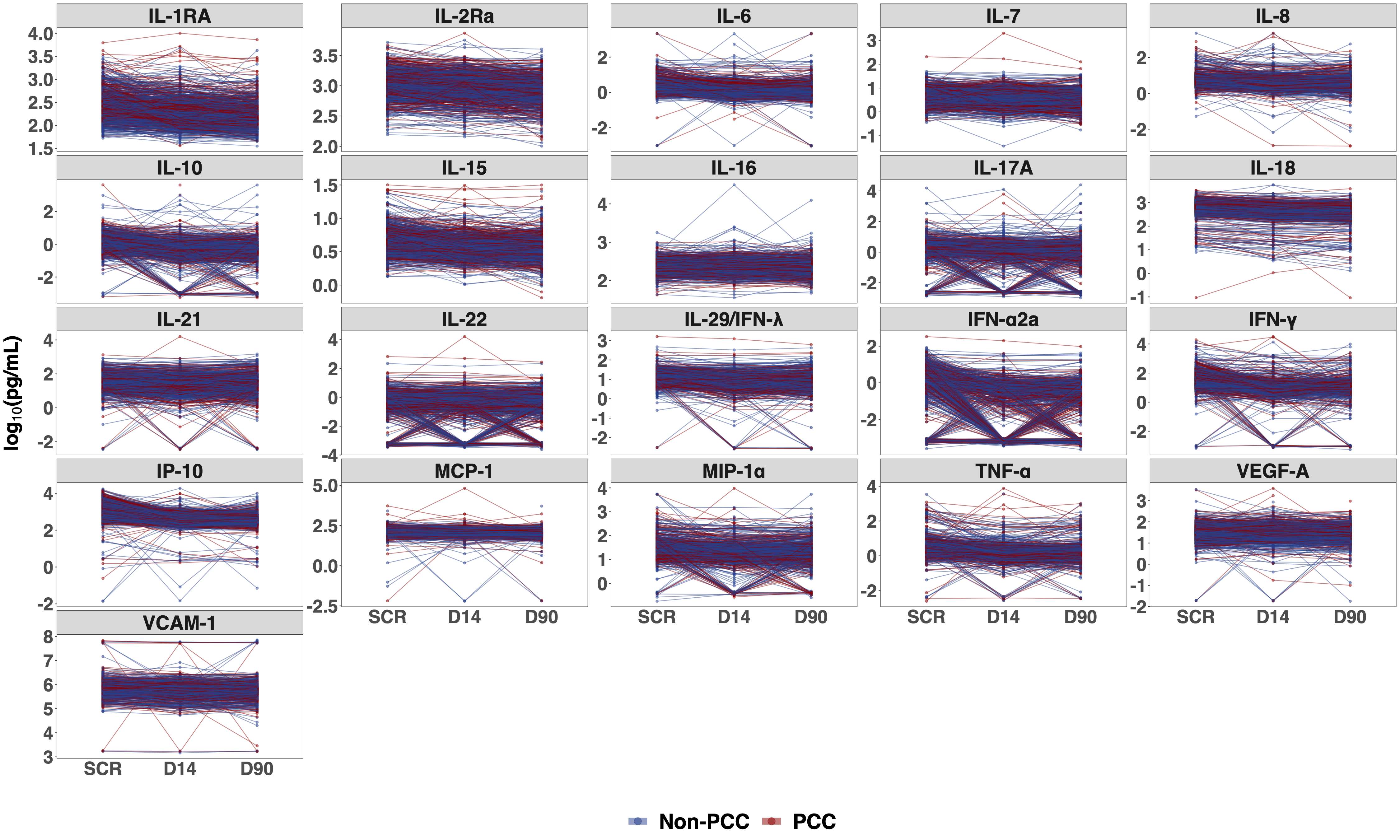

**Supplemental Figure 2. Comparison of cytokines between PCC and non-PCC at screening, day 14 and day 90**

Abbreviation: PCC: Post COVID 19 condition. SCR: screening visit. D14: Day 14 visit. D90: day 90 visit.

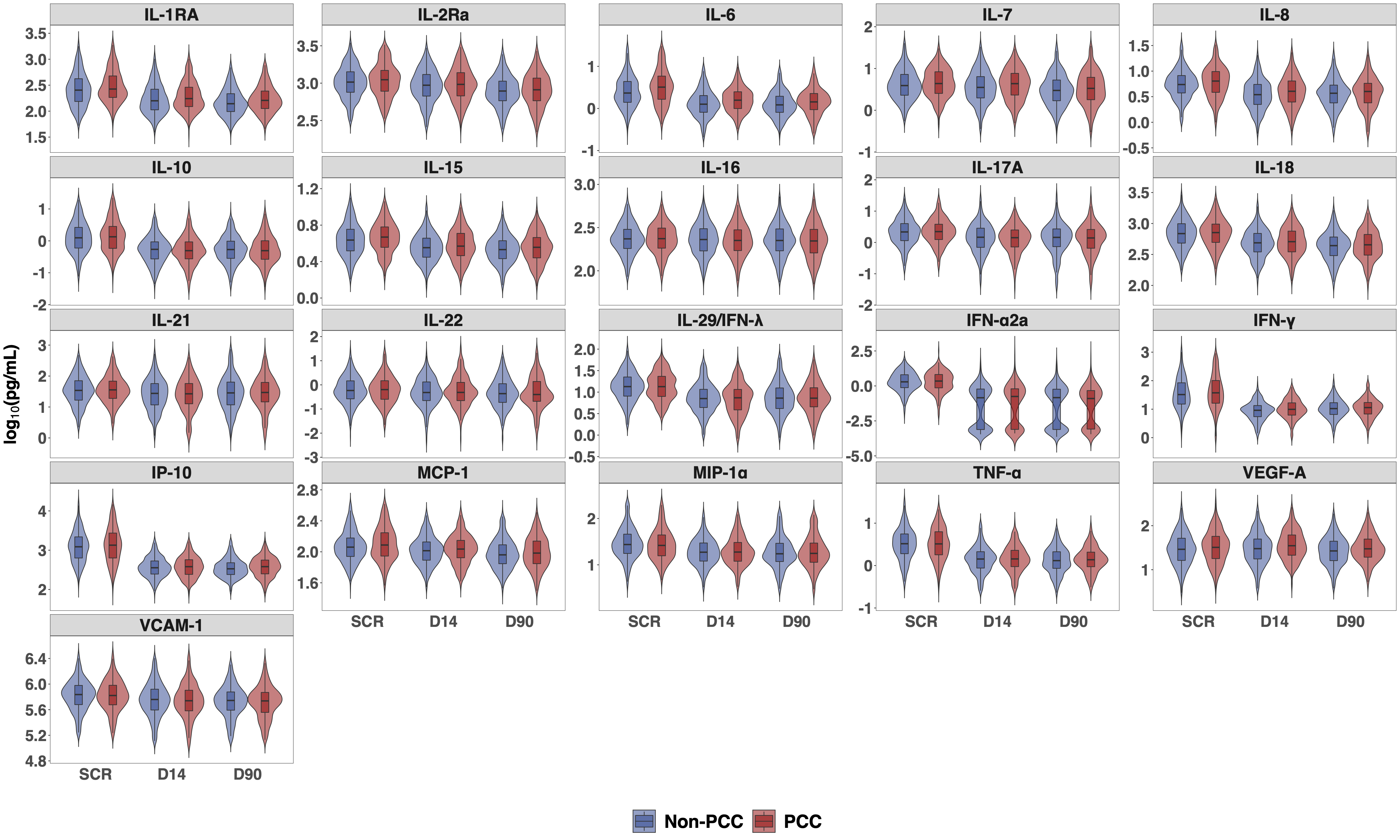

**Supplemental Table 5. Multivariable logistic regression between log_10_ cytokine levels at screening and PCC**

| Analyte | OR (95% CI) | P value |
| --- | --- | --- |
| IL-1RA | 1.44 (0.83 - 2.50) | 0.189 |
| IL-2Ra | 1.43 (0.67 - 3.08) | 0.361 |
| IL-6 | 1.59 (1.02 - 2.47) | **0.039** |
| IL-7 | 0.95 (0.59 - 1.51) | 0.819 |
| IL-8 | 0.97 (0.52 - 1.82) | 0.936 |
| IL-10 | 1.02 (0.73 - 1.42) | 0.911 |
| IL-15 | 2.43 (0.92 - 6.45) | 0.073 |
| IL-16 | 1.00 (0.39 - 2.58) | 0.996 |
| IL-17A | 0.96 (0.64 - 1.44) | 0.851 |
| IL-18 | 1.04 (0.50 - 2.15) | 0.921 |
| IL-21 | 1.11 (0.78 - 1.57) | 0.559 |
| IL-22 | 1.02 (0.76 - 1.38) | 0.888 |
| IL-29/IFN lambda | 1.13 (0.69 - 1.86) | 0.629 |
| IFN-α2a | 0.95 (0.74 - 1.22) | 0.675 |
| IFN-γ | 1.06 (0.79 - 1.41) | 0.712 |
| IP-10 | 0.92 (0.60 - 1.40) | 0.685 |
| MCP-1 | 1.44 (0.64 - 3.25) | 0.374 |
| MIP-1α | 0.74 (0.45 - 1.20) | 0.218 |
| TNF-α | 0.84 (0.54 - 1.33) | 0.462 |
| VEGF-A | 1.02 (0.65 - 1.60) | 0.919 |
| VCAM-1 | 0.90 (0.47 - 1.73) | 0.750 |

Abbreviation: PCC: Post COVID 19 condition. OR: Odds Ratio. CI: Confidence Interval.

Note: Each cytokine was modeled in separate logistic regression. OR represents per unit increase in log_10_ pg/mL for each cytokine. Multivariable model adjusted for baseline age (continuous), sex (male vs female), race (white vs non-white), BMI (<30 vs ≥30), vaccine status (partially or fully vs none), diabetes (no vs yes), and plasma transfused (control plasma vs CCP>5 days since illness vs CCP≤5 days since illness). P<0.05 were highlighted in bold. Except for IL-6, no p value was statistically significant after adjusting for multiple comparison using Benjamini-Hochberg correction with a false discovery rate of 0.05.

**Supplemental Table 6a. Logistic regression between demographic and clinical factors and PCC among the people in the original trial and have day 90 symptom data**

|  | OR (95% CI), n = 1061 | |
| --- | --- | --- |
| Analyte | Univariate | Multivariate |
| Age, scaled to 10 years | **1.26 (1.15, 1.38)** | **1.31 (1.18, 1.45)** |
| Sex |  |  |
| Male | Ref. | Ref. |
| Female | **2.12 (1.62, 2.78)** | **2.31 (1.74, 3.09)** |
| Race |  |  |
| White | Ref. | Ref. |
| Non-white | 0.77 (0.55, 1.06) | 0.73 (0.51, 1.04) |
| BMI |  |  |
| <30 | Ref. | Ref. |
| ≥30 | **1.44 (1.10, 1.88)** | 1.29 (0.97, 1.72) |
| Vaccine Status |  |  |
| None | Ref. | Ref. |
| Partial or full | 0.81 (0.58, 1.12) | 0.86 (0.61, 1.22) |
| Diabetes |  |  |
| No | Ref. | Ref. |
| Yes | 1.49 (0.96, 2.29) | 1.25 (0.76, 2.02) |
| Plasma transfused |  |  |
| Control plasma | Ref. | Ref. |
| CCP>5 days since illness | 1.04 (0.77, 1.40) | 1.18 (0.85, 1.62) |
| CCP≤5 days since illness | 0.79 (0.57, 1.11) | 0.90 (0.63, 1.28) |

**Supplemental Table 6b. Logistic regression between demographic and clinical factors and PCC among the people having day 90 symptom data and transfused with CCP in the original trial**

|  | OR (95% CI), n = 533 | |
| --- | --- | --- |
| Analyte | Univariate | Multivariate |
| Age, scaled to 10 years | **1.27 (1.12, 1.45)** | 1.33 (1.15, 1.53) |
| Sex |  |  |
| Male | Ref. | Ref. |
| Female | **1.69 (1.17, 2.46)** | **2.08 (1.40, 3.14)** |
| Race |  |  |
| White | Ref. | Ref. |
| Non-white | 0.72 (0.45, 1.13) | 0.67 (0.40, 1.08) |
| BMI |  |  |
| <30 | Ref. | Ref. |
| ≥30 | **1.51 (1.03, 2.22)** | 1.48 (0.98, 2.21) |
| Vaccine Status |  |  |
| None | Ref. | Ref. |
| Partially or fully | 0.73 (0.44, 1.16) | 0.76 (0.45, 1.23) |
| Diabetes |  |  |
| No | Ref. | Ref. |
| Yes | 1.27 (0.67, 2.32) | 1.02 (0.51, 2.03) |
| Plasma transfused |  |  |
| CCP>5 days since symptom onset | Ref. | Ref. |
| CCP≤5 days since symptom onset | 0.76 (0.53, 1.10) | 0.76 (0.51, 1.12) |

Abbreviation: PCC: Post COVID 19 condition. OR: Odds Ratio. CI: Confidence Interval.

Note: of the 1061 patients with 90 day data, 355 (33%) had PCC. Of those 355, 181 were in the control treatment group, 106 had CCP>5 days and 68 had CP≤ 5 days. Of the 706 participants (67%) who did not have PCC, 347 were in the control treatment group, 195 had CCP>5 and 164 had CCP ≤ 5 days after symptom onset.
